# Temporal Immune Effects of Oral Ketamine on PTSD: Transcriptomic Evidence of Short-Term Inflammation Suppression and Long-Term Immune Remodelling

**DOI:** 10.1101/2025.05.26.25328370

**Authors:** Nathan J. Wellington, Bonnie L. Quigley, Ana P. Bouças, Megan Dutton, Adem T. Can, Jim Lagopoulos, Anna V. Kuballa

## Abstract

Ketamine’s rapid acting symptom relief make it a promising intervention for PTSD. However, the mechanisms driving its long-term efficacy over weeks and months remain poorly understood. This study investigated the short-and long-term impacts on gene expression of a six-week subanesthetic oral ketamine trial in 23 PTSD participants (9 males, 14 females). Peripheral Blood Mononuclear Cells (PBMCs) were collected at baseline, one week (short-term), and four weeks (long-term) post oral ketamine for RNA sequencing and transcriptome analysis. Differential expression analysis identified substantial and persistent transcriptomic changes over time, with 533 genes upregulated and 621 downregulated across timepoints. Notably, there was a 37% increase in differential gene expression between the short-and long-term responses, accompanied by a 6.5-fold rise in expression magnitude and an 8.8-fold enhancement in pathway activity. Pathway analysis emphasised critical immune and inflammatory pathways that appear to be modulated by ketamine, including interferon alpha/beta signalling (*z =* 4), IL-17 signalling pathway (*z =* 3.36), and cytokine storm signalling (*z =* 4.26), neutrophil degranulation (z = 6.0) and antimicrobial peptide signalling (*z =* 1.63) which differed across timepoints. The findings suggest a transition from short-term inflammation suppression and antimicrobial activity to long-term sustained immune regulation, inflammation remodulation and tissue repair. Key cytokines, chemokines, interferons and antimicrobial peptides included, IL-6, IL-1β, IFI27, IL-10 signalling, CXCL8, SOCS1/3 and CAMP which represent central regulators of immune and inflammatory pathways. These molecular changes offer novel insights into the short-and long-term therapeutic potential of ketamine for PTSD and highlight avenues for precision psychiatry and individualised maintenance therapy to prevent relapse.

## 1 Introduction

Post-traumatic stress disorder (PTSD) is a severe and chronic psychiatric condition that can develop following exposure to traumatic events such as war, natural disasters, or interpersonal violence [1]. Despite the availability of pharmacological and psychotherapeutic interventions, a large subset of individuals with PTSD remain resistant to treatment, highlighting the need for novel therapeutic strategies that target the core neurobiological underpinnings of the disorder [2].

In recent years, ketamine has garnered attention as a promising treatment for PTSD, particularly due to its rapid and sustained antidepressant effects [3–7]. Ketamine’s ability to disrupt traumatic memory reconsolidation is also rooted in its action as an NMDA receptor antagonist [8]. By interfering with the calcium influx and intracellular signalling, subanesthetic doses of ketamine have been shown to prevent the memory from stabilising effectively weakening the trace memory often within hours of administration [9, 10]. While the clinical efficacy of ketamine in ameliorating PTSD symptoms has been demonstrated in several studies, the precise molecular mechanisms underlying its therapeutic effects remain poorly understood [7, 11–14].

PTSD has also been linked to chronic levels of inflammation, including pro-inflammatory cytokines, such as interleukin-1β (IL-1β), interleukin-6 (IL-6), and tumour necrosis factor-α (TNF-α), as well as acute-phase proteins, such as C-reactive protein (CRP), in the blood [15, 16]. In preclinical models ketamine has been found to modulate these inflammatory responses by supressing microglia and inhibiting the release of these cytokines [17]. An *in vitro* study on human neutrophils found that ketamine inhibits the up-regulation of CD18, shedding of CD62L, oxygen radical production, and reduces IL-6 production in endotoxin-stimulated blood, suggesting its potential role in modulating immune responses during inflammatory conditions like sepsis [18]. Another recent study of healthy males found that ketamine inhibited transcription factors NF-κB and AP-1. These factors are crucial in regulating the production of pro-inflammatory mediators, which are involved in inflammatory responses [19]. It was also found that ketamine significantly inhibited phagocytosis and bacterial activity through the suppression of polymorphonuclear leucocytes (PMNL) *in vitro* [20].

By simultaneously reducing inflammation and promoting neuroplasticity, ketamine creates a dual mechanism for alleviating PTSD symptoms. However, little is known about whether these inflammatory or plasticity changes are sustained over longer periods following ketamine treatment or whether they revert back to baseline levels. Ketamine’s long-lasting impact on gene expression is particularly confounded by a heavy reliance on preclinical studies and rodent model research [21–23]. Chronic subanesthetic dosing of ketamine has been employed in rat-and mouse-model preclinical and clinical studies to mimic the therapeutic protocols used in humans, where repeated dosing over several weeks can result in prolonged symptom relief [24–26]. This pharmacological approach, when combined with psychotherapy, has been regarded as highly effective, delivering immediate therapeutic benefits [21, 27–30]. However, there is little research regarding PTSD relapse rates post ketamine treatment, or evidence based therapeutic models for maintenance protocols for ongoing ketamine treatment for PTSD, with the bulk of the research conducted in preclinical environments [21, 31–34].

It is hypothesised that ketamine’s therapeutic effects on PTSD are mediated, at least in part, by its longer-term influence on the transcriptome, leading to persistent changes in gene expression that support neuroplasticity, immune modulation and stress resilience. The transcriptome, representing the complete set of RNA transcripts produced by the genome, is a dynamic reflection of cellular activity and can be modulated by both genetic and epigenetic factors [35]. In this study, we sought to advance our understanding of cellular responses to ketamine by investigating both the short-term and more sustained effects of an oral subanesthetic ketamine treatment on the transcriptome in PTSD. The findings from this study have the potential to progress the development of sustained psychopharmacological therapies for PTSD, ultimately improving outcomes for individuals suffering from this debilitating condition.

## 2 Methods

### 2.1 Study Design

This study builds upon the Oral ketamine treatment of PTSD (OKTOP), which was conducted between July 2021 and January 2024 under the ethical oversight of the Prince Charles Hospital (PCH) Human Research Committee (ethics approval number HREC/18/QPCH/288) and the University of the Sunshine Coast (USC; ethics approval number A181190) and registered with the Clinical Trials Registry of Australia and New Zealand (ACTRN12618001965291) [36]. Genetic analysis and reporting were part of the Genetic Biomarkers of Ketamine on PTSD (GBOK) project, with ethics approvals from PCH (HREC/18/QPCH/288) and UniSC (S211655). The OKTOP trial was an open-label, dose-ranging clinical investigation designed to evaluate the effectiveness, feasibility, and tolerability of oral ketamine in treating PTSD. The study spanned 10 weeks, comprising six weeks of active treatment followed by two follow-up assessments (**Fig.1**).

**Fig. 1:**
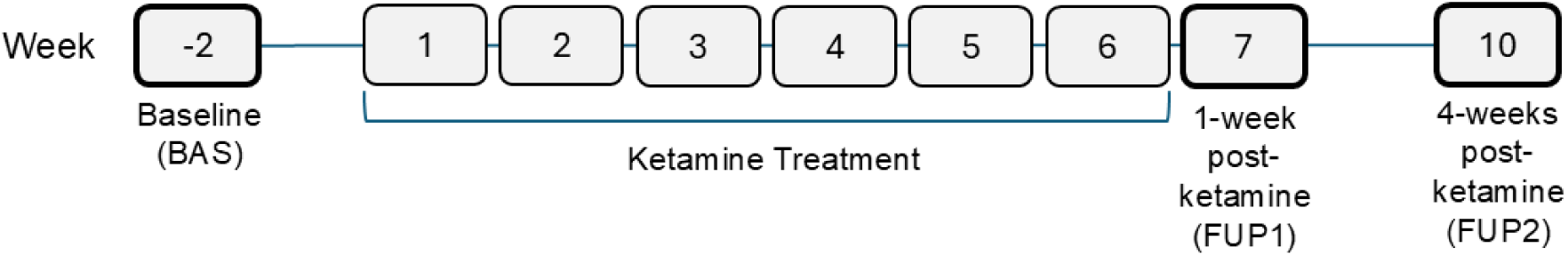
An overview of the OKTOP Study Design. Study timeline illustrating the experimental design for ketamine treatment and follow-up assessments. Baseline (BAS) was established 2 weeks prior to treatment commencement, followed by a 6-week ketamine treatment (one dose per week) spanning weeks 1 to 6. Post-treatment assessments were conducted at week 7 (1-7 days post ketamine, FUP1) and week 10 (28-32 days post ketamine, FUP2). Blood samples were collected at BAS, FUP1 and FUP2 for this study.

Research blood samples were collected at the baseline assessment performed 2 weeks prior to treatment commencement (BAS), 1-7 days post-ketamine treatment (FUP1), and 28-32 days post-ketamine treatment (FUP2). During the intervention, participants received a single weekly dose of oral ketamine at subanaesthetic levels, administered under the supervision of a consultant psychiatrist. The starting dose was 0.5 mg/kg, with weekly increases ranging from 0.1 mg to 0.5 mg per kg, depending on individual tolerance. Dose titration continued until the maximum tolerated level was attained, capped at a maximum dose of 3.0 mg/kg. The average dose across participants was 1.4 mg/kg per week (SD = 0.37) (**Table 1**).

**Table 1:**
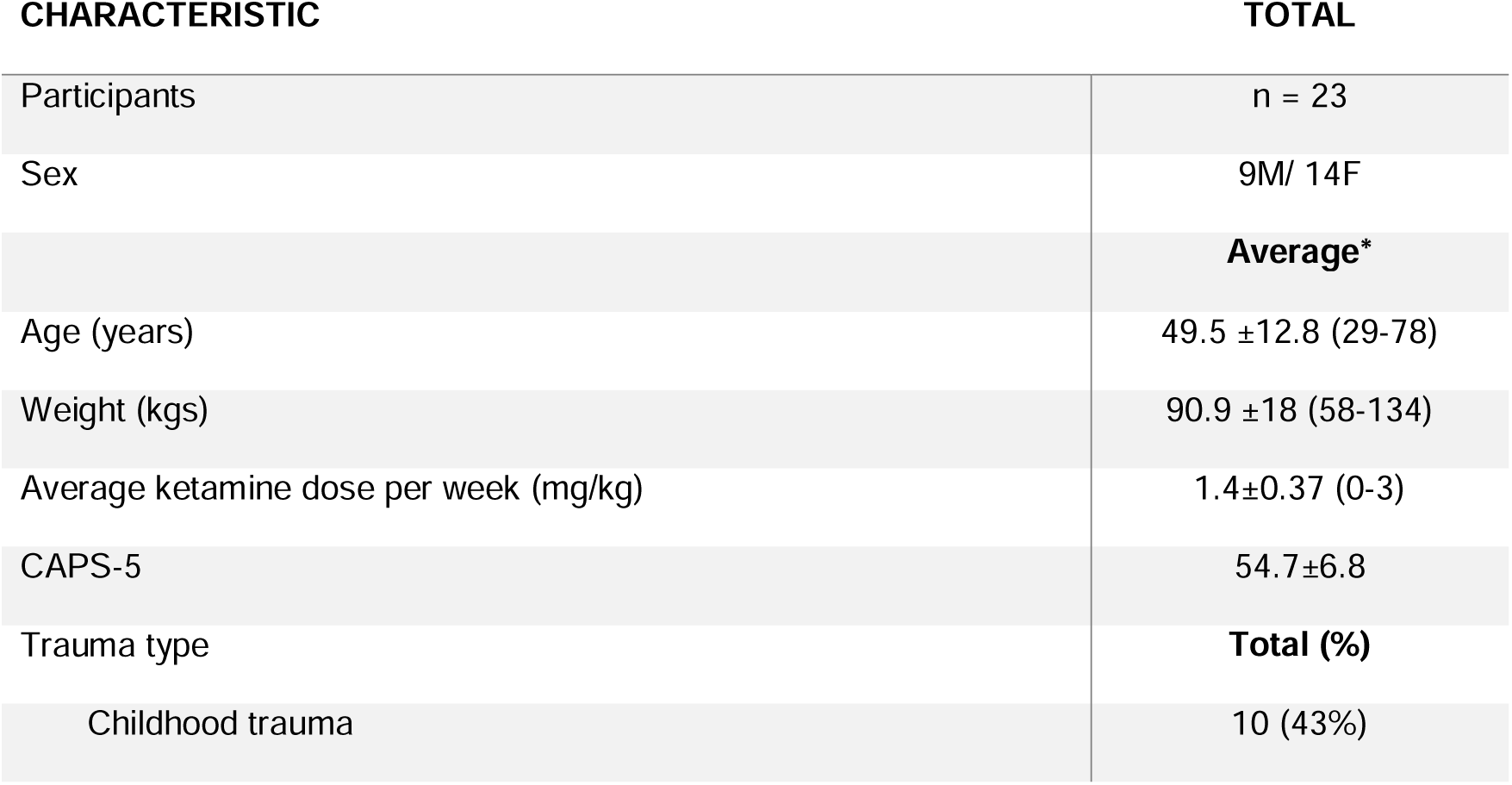
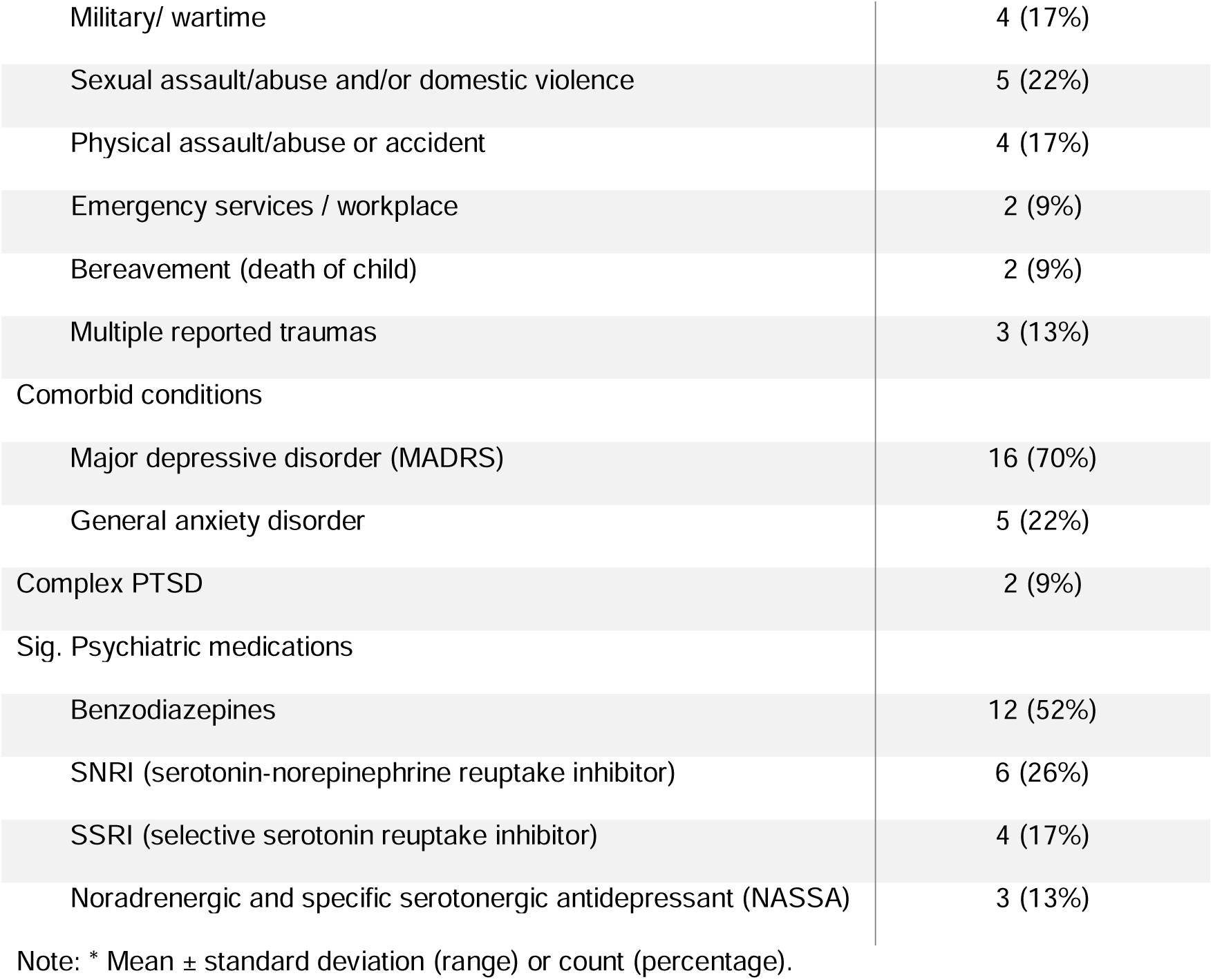
Participant characteristics including clinical diagnoses, trauma type, comorbid conditions, and medications.

### 2.2 Participants

A total of 23 PTSD participants (9M/14F) that met the OKTOP inclusion criteria [36] were included in this study, all of whom were Caucasian, with ages ranging from 29 to 78 years (M = 49.5, SD = 12.8) (**Table 1**). Body weight varied between 58 and 134 kg (M = 90.9, SD = 18). Baseline assessments were conducted within 14 days prior to starting treatment and included a physical examination, urinalysis, medical history review, and both research and clinical blood tests. Participants were diagnosed using the Clinician-Administered PTSD Scale for DSM-5 (CAPS-5), with a PTSD diagnosis defined as meeting Criteria B, C, D, and E for more than 1 month with a score ≥ 33 and severe PTSD defined as above with a score ≥ 50. The cohort had an average CAPS-5 score of 54.7 (SD = 6.8) [37]. Secondary assessments were conducted using the Montgomery–Åsberg Depression Rating Scale (MADRS) and the Depression, Anxiety, and Stress Scale (DASS-21) [38, 39]. Additionally, 70% of participants were also diagnosed with MDD (**Table 1**).

Further self-assessments, including the PTSD Checklist for DSM-5 (PCL-5) and the World Health Organisation (WHO) quality of life measures, were performed throughout the trial and at post-intervention follow-up to monitor participants’ responses [40, 41]. Only individuals who attended all ketamine sessions and provided at least two blood samples (BAS and a comparative time point: FUP1 or FUP2) were included in the final analysis.

### 2.3 Blood Collection

Non-fasting whole blood samples were drawn from all participants for analysis at BAS, FUP1 and FUP2, with 50-70 ml of whole blood collected by a phlebotomist and processed into aliquots of whole blood, serum, plasma, and peripheral blood mononuclear cells (PBMCs). Further > 87% of all samples were drawn and processed for between 9-10:30am controlling for potential diurnal variation. PBMCs were resuspended in 600 μL RNeasy lysis buffer (Qiagen) and 6 μL of beta-mercaptoethanol to preserve RNA integrity and stored at-80°C.

### 2.4 RNA-Seq Acquisition

A total of 61 PBMC samples suspended in lysis buffer (BAS *=* 23, FUP1 = 19 and FUP2 *=* 19) (GSE288174) were transported on dry ice to the Australian Genome Research Facility (AGRF) for RNA sequencing analysis. RNA was extracted via the RNeasy mini kit (Qiagen) as per manufacturer’s instructions for cells. Gene expression analysis was conducted on a NovaSeq utilising the Illumina RNA-Seq 20 million paired end sequencing protocol. Image analysis was carried out in real-time using NovaSeq Control Software (NCS) v1.2.0.28691 and Real-Time Analysis (RTA) v4.6.7 on the instrument computer. The Illumina DRAGEN BCL Convert pipeline (07.021.645.4.0.3) was used to produce sequence data. The data underwent quality control, trimming, alignment, annotation, and differential expression analysis using CLC Genomics Workbench 25.0 (Qiagen). Samples that were missing at collection, did not meet a RIN value of ≤9.6 for sequencing, or did not pass quality control (*n =* 9) were accounted for in co-weighting for clustering within data analysis. Reads were mapped to *Homo sapiens* GRCh38.p14, RNA-seq and CDS databases for annotation.

### 2.5 Network and Pathway Analysis

Network and pathway analysis was conducted utilising Ingenuity Pathway Analysis 24.0.1 (IPA) (Qiagen). Gene enrichment and ontology was conducted utilising the Gene Ontology knowledgebase [42]. Canonical pathways were calculated using the right tailed fisher’s exact testing model. Graphics were generated through both platforms and complemented with SRPlot [43]. RNA seq data was imported into IPA utilising RPKM, fold change, log fold change and FDR p-value (*q*-value) data and mapped to HUGO Gene Nomenclature Committee (HGNC) and RefSeq release 227 databases. The data was analysed through the core analysis function, considering co-weighting for samples not included, and applying a *q*-value <0.05 and interpreted through fold change ≤-1.3 or ≥ 1.3. Additional RNA-Seq data analysis was performed in R-Studio 2024.04.2 build 764 and utilising multiple linear regression analysis to calculate covariate and RQdeltaCT, DESeq2 and clusterProfiler to calculate the transcriptomic and canonical pathway magnitude of change through Mann-Whitney U nonparametric testing [44–47].

## 3 Results

### 3.1 Clinical Covariates

To control for confounding factors influencing the clinical response to ketamine in PTSD a multiple linear regression model was performed with clinician-rated PTSD severity (CAPS-5) as the dependent variable and sex, age, body weight, self-reported PTSD symptoms (PCL-5), and depressive symptoms (MADRS) as covariates. The model accounted for 55% of the variance in CAPS-5 scores (R² = 0.55, adjusted R² = 0.42; F(5,17) = 4.12, *p* = 0.012), suggesting a moderate explanatory power for baseline symptom severity and individual characteristics in shaping ketamine response. Among the predictors, PCL-5 (β = 0.25, *p* = 0.061) and MADRS (β = 0.33, *p* = 0.097) showed trend-level associations with the BAS CAPS-5, indicating that higher baseline PTSD and depressive symptom burden may moderate by the therapeutic effects of ketamine. In contrast, sex, age, and weight had no significant influence on scores post-treatment. These results imply that while demographic factors exert minimal impact, individual symptom profiles, particularly self-reported PTSD and depression may shape the degree of clinical benefit derived from ketamine. These variables were therefore included as covariates in downstream gene expression analyses to control for confounding influences when evaluating the molecular correlates of ketamine’s therapeutic effects in PTSD (*N = 23; R² = 0.55, Adjusted R² = 0.42; F(5,17) = 4.12, p = 0.012*). (See **Table 2**).

**Table 2.**
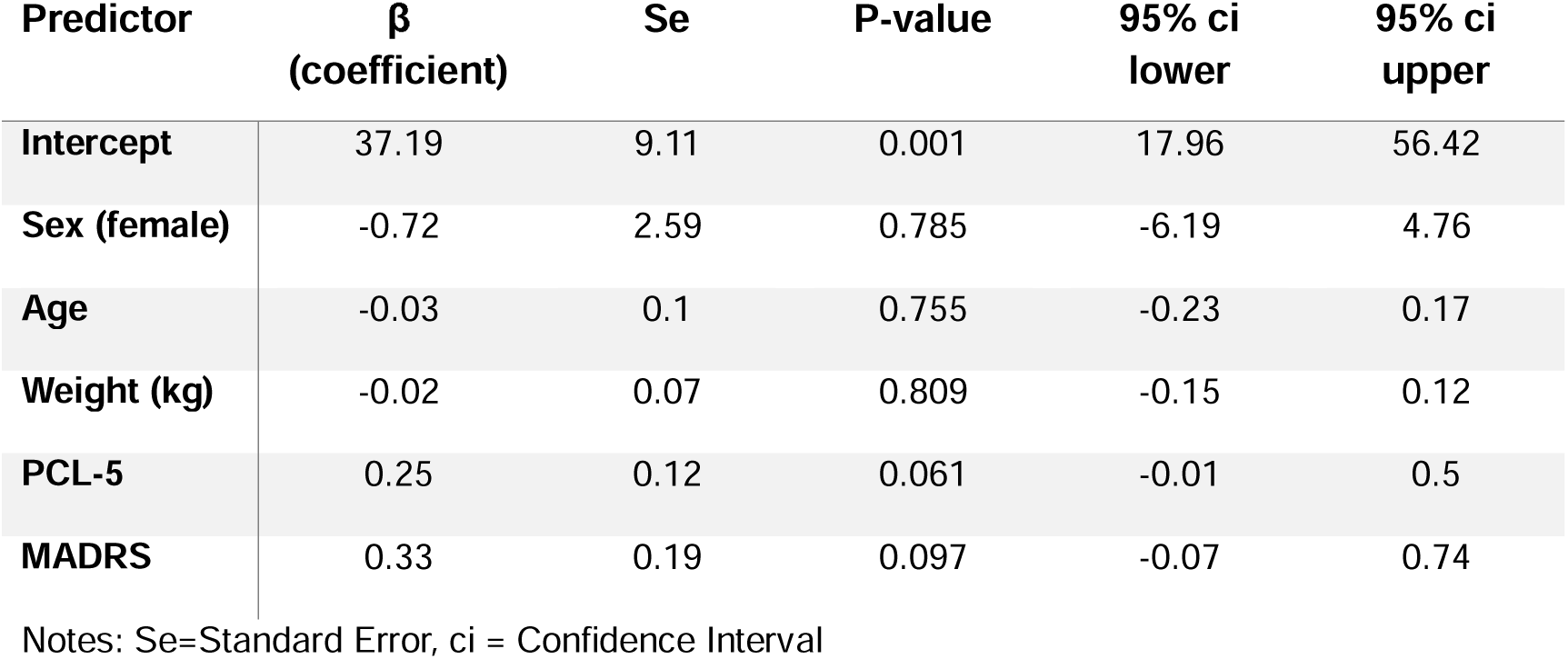
Linear Regression Predicting Treatment Outcome.

### 3.2 Transcriptomic changes across Timepoints

Accounting for these covariates, differential gene expression analysis was performed comparing BAS with FUP1 and FUP2 to determine the longitudinal gene expression changes post ketamine treatment. RNA-seq analysis identified a total of 59,116 transcripts, including long noncoding RNAs, complementary RNAs and mitochondrial RNAs (GSE288174). Data analysis accounting for a *q*-value of ≤ 0.05 and expression fold changes <-1.3 and >1.3, identified 1028 genes that were significantly differentially expressed, from BAS across FUP1 and FUP2 with 533 up-regulated and 621 down-regulated genes (**Fig.2**). A total of 126 differentially expressed transcripts were common to both FUP1 and FUP2. Of these, 66 were down-regulated in FUP1, 60 of these continued to be down-regulated in FUP2. Additionally, 60 genes were up-regulated in FUP1, 59 of these continued to be up-regulated at FUP 2 while 7 genes changed expression direction. (**Fig.2, Supplementary Table 3**).

**Fig. 2:**
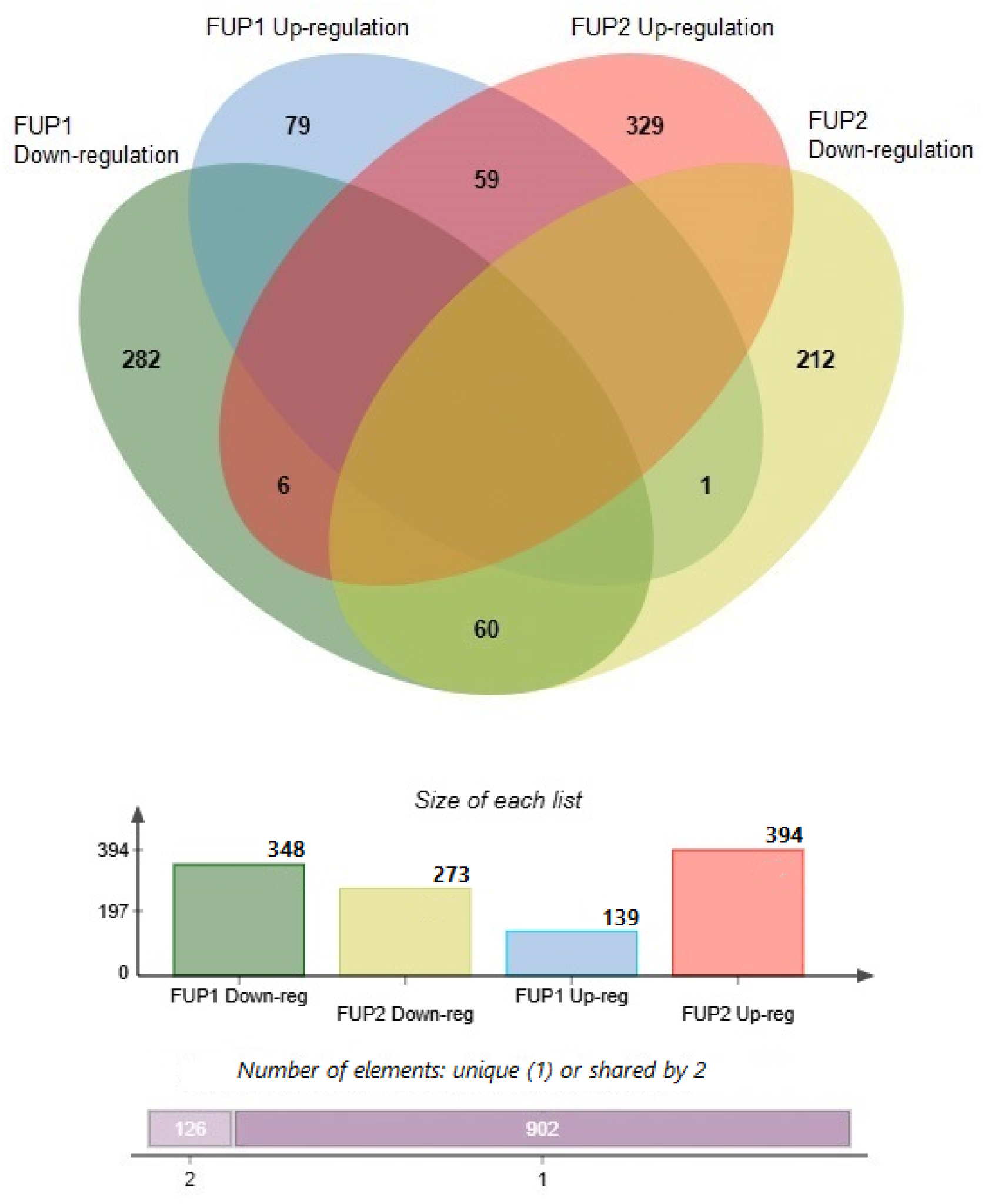
Venn diagram showing the overlap of up-regulated and down-regulated genes comparing BAS to both FUP1 and FUP2 timepoints. Each coloured circle represents a gene set: FUP1 down-regulation (green), FUP1 up-regulation (blue), FUP2 up-regulation (red), and FUP2 down-regulation (yellow). The numbers in the intersections represent shared genes, while non-overlapping regions indicate condition-specific genes. Below the Venn diagram, a bar graph displays the total number of genes identified within each timepoint, and a summary bar indicates the number of unique (1) and shared (2) genes across the conditions.

**Fig. 3:**
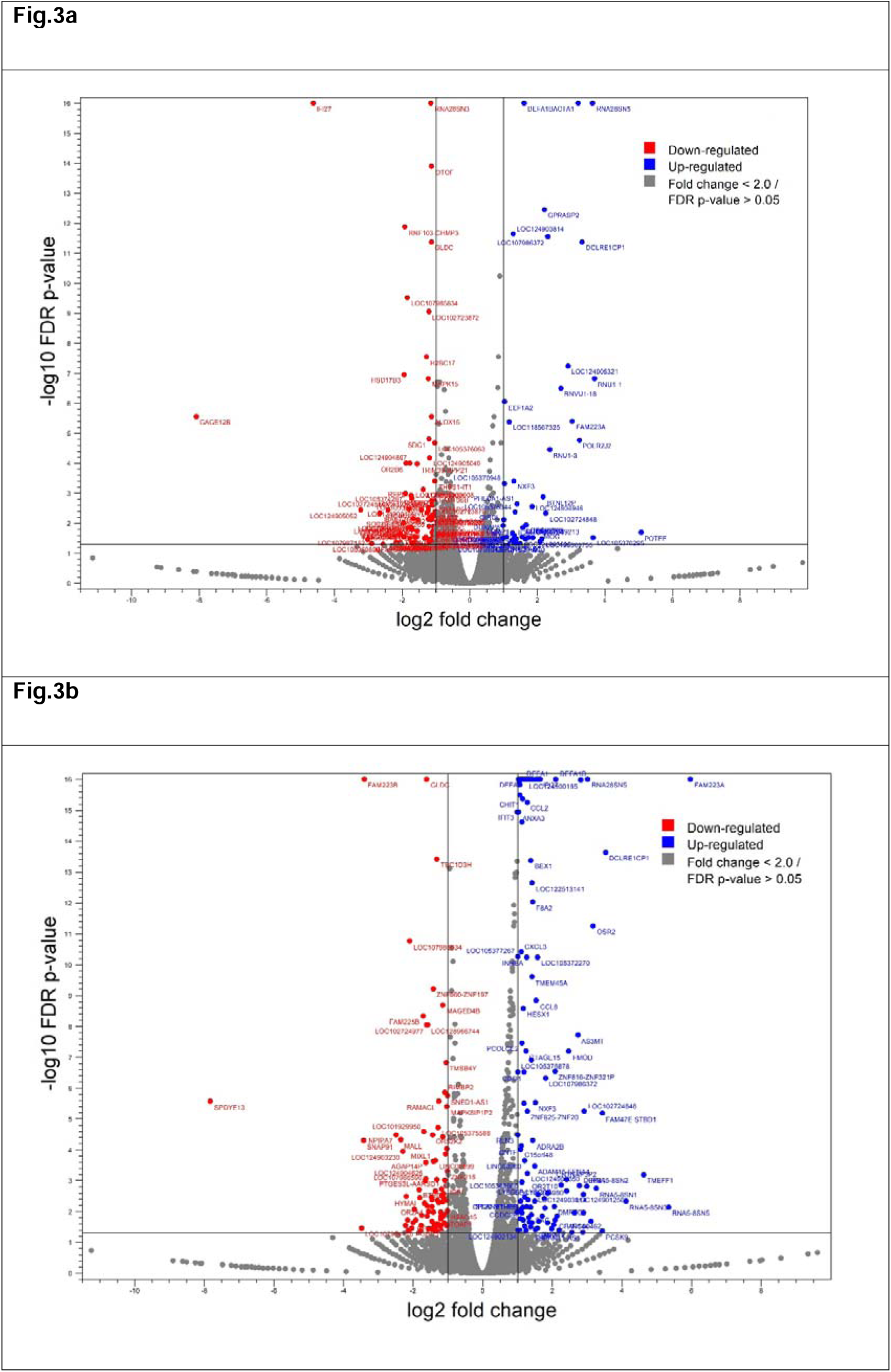
Volcano plots displaying differentially expressed genes comparing short term (a) and long term (b) fold changes (log2 scale) and statistical significance (-log10 FDR p-value) to ketamine therapy. The x-axis represents the log_2_ fold change, where values to the left (red) indicate down-regulation and values to the right (blue) indicate up-regulation. The y-axis shows the-log10 transformed FDR-adjusted p-values, with higher points indicating greater statistical significance. Genes meeting the significance threshold (FDR p-value ≤ 0.05 and absolute fold change ≥ 2) are coloured Key genes with significant expression changes are labelled. The horizontal line represents the FDR threshold, and the vertical lines indicate the fold change cutoff values (±2).

**Fig. 4:**
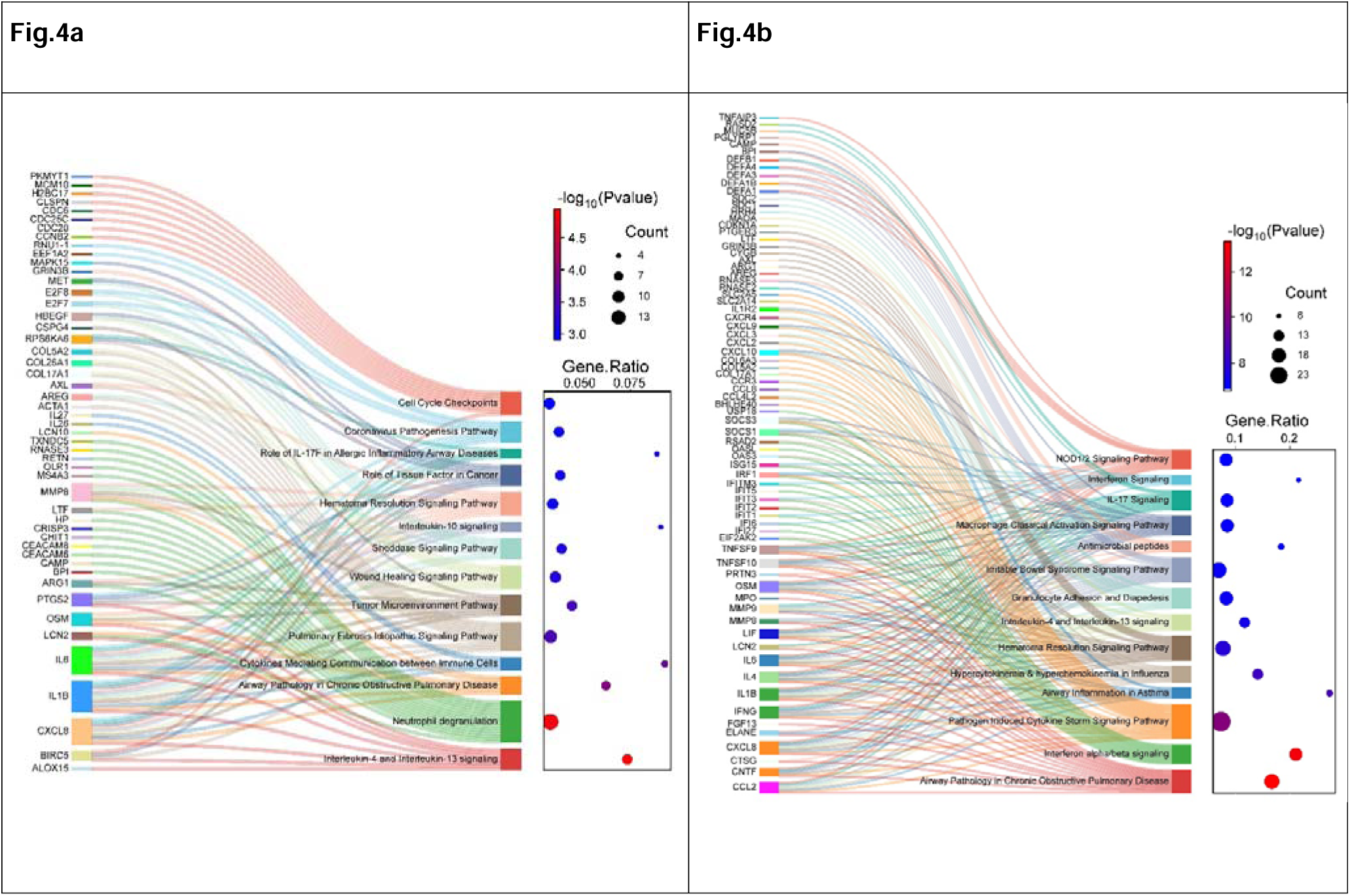
Canonical pathways for the short-term (4a) and long-term (4b) effects of ketamine treatment. Each Sankey diagram highlights the connections between individual differentially expressed genes (left column) and their respective enriched canonical pathways (right column), as identified by pathway analysis. Each gene is linked to one or more pathways, with pathways categorised based on their biological relevance, such as “immune signalling”, “neutrophil degranulation” and “wound healing”. The bubble plot on the right represents the statistical significance (-log10(p-value)) and gene ratio for each pathway. The size of the bubbles indicates the number of genes involved in each pathway, while the colour gradient reflects the level of significance (red = higher significance). Pathways such as “cell cycle checkpoints” and “neutrophil degranulation” show high levels of enrichment with multiple contributing genes.

When determining the differences in gene expression across the time points, we found the short-term effects of ketamine between BAS and FUP1 (1-week post treatment) revealed 487 differentially expressed genes, of which 348 were down-regulated and 139 were up-regulated (**Fig.3a, & Supplementary Table 1**). Alternatively, the long-term effects of ketamine between BAS and FUP2 (4-weeks post treatment) identified 667 differentially expressed genes, of which 273 were down-regulated and 394 were up-regulated (**Fig.3b, Supplementary Table 2**).

The changes in the number of genes differentially expressed between time points increased by 36.9% in the longer term (BAS to FUP2) compared to the short-term response (BAS to FUP1) following ketamine treatment (**Fig.2, 3a & 3b**). Additionally, when comparing the directionality of genes, up-regulation increased by 238.4% from FUP1 to FUP 2 (*n* = 255) whilst there was a 28.9% decrease in down-regulated genes at FUP2 (*n* = 134) (**Fig.2, 3a & 3b**). Finally, when examining the fold change magnitude between time points, the average magnitude of change at FUP2 was 6.5 times that of FUP1, indicating a substantial increase in gene expression activity (*U*=144368, *p*=0.0012).

### 3.3 Network Changes across Timepoints

Mapping these genes to canonical pathways revealed several key functional mechanisms that may be affected by ketamine treatment. The short-term effect of ketamine was broadly grouped into three themes; the immune system, cytokine signalling and cellular stress (**Fig.4a, Supplementary Table 4**). The canonical pathways most influenced by ketamine at FUP1 include antimicrobial activity against bacteria (z = 1.63), neutrophil degranulation (z = 3), tumour microenvironment (z = 2.21), wound healing signalling (z = 1), interleukin-10 (IL-10) signalling (z = 2), interleukin-4 (IL-4) and interleukin-13 (IL-13) signalling (**Fig.4a**).

The top canonical pathway clusters affected by ketamine treatment 4 weeks post ketamine therapy (FUP2, long-term) indicate that there was continued influence on the immune system, cytokine signalling and cellular stress with additional focus on inflammation, neurotransmitter and nervous system signalling (**Fig.4b,**

**Supplementary Table 5**). The key enriched long-term pathways showed increases in the activation of immune related pathways such as; neutrophil degranulation (which doubled in activation from FUP1) (*z* = 6), wound healing signalling (with a 3-fold increase from FUP1) (*z* = 3.3), antimicrobial peptides (which almost doubled from FUP1) (*z* = 3), and IL-10 signalling (which increased by 40% from FUP1) (*z* = 2.8). Other significantly active pathways included interferon alpha/beta signalling (*z* = 4), pathogen induced cytokine storm signalling pathway (*z* = 4.26), IL-4 and IL-13 signalling (*z* = 1.94), macrophage classical activation signalling pathway (*z* = 2.84), IL-17 signalling (*z* = 3.36) and interferon signalling (*z* = 3.35) (**Fig.4b, Supplementary Table 5**). When comparing the *z*-scores of canonical pathways between FUP1 and FUP2, the overall *z*-scores in FUP2 showed an elevation in activity of approximately 8.8-fold, indicating a significant increase in pathway magnitude 4 weeks post ketamine therapy (*U* = 20363, *p* = 3.63 x 10^-179^). This also reflected the increase in expression activity and the fold change values observed in many other genes (**Supplementary Table 5**).

## 4 Discussion

This is the first longitudinal transcriptomic analysis on the molecular effects of ketamine in the treatment of PTSD, in which the short-term and long-term effects of six-week oral subanesthetic ketamine administration were explored. The findings in this study reveal significant and sustained changes to gene expression in pathways related to long-term immune response and inflammation. This highlights ketamine’s dual mechanism of action affecting both immunomodulation and neuroplasticity and provides new insights into biological processes and potential targets for future PTSD psychopharmacology.

The differential expression profiles of PTSD patients administered ketamine reveal dynamic magnitude of expression changes over time, with a significant 36.9% increase in differentially expressed genes between FUP1 and FUP2. Furthermore, the fold change of expression values also exhibited an overall increase in magnitude of over 6.5 times that from FUP1 to FUP2. When comparing the *z*-scores of canonical pathways between FUP1 and FUP2, the overall *z*-scores in FUP2 showed an elevation of approximately 8.8-fold in magnitude, indicating a substantial increase in pathway activity 4 weeks post ketamine therapy. These findings underscore the significant longitudinal effects of ketamine on gene expression and pathway activation.

### 4.1 Modulation of Pro-Inflammatory Pathways

This study demonstrates that ketamine significantly alters gene expression related to inflammatory signalling, with both short-and long-term effects observed at FUP1 and FUP2, respectively in key pathways such as cytokine signalling (z = 4.26) and IL-4 and IL-13 signalling (z = 1.94). These pathways were activated at FUP1 but exhibited sustained modulation at FUP2, indicating ketamine’s ability to regulate inflammatory responses over time. Ketamine was also found to downregulate pro-inflammatory cytokines, including IL-1β (*q* = 2.92 × 10⁻ ²) and IL-6 (*q* = 0.0008), as well as the chemokine CCL2 (*q* = 5.56 × 10⁻¹) and the interleukin receptor IL1-R2 (*q* = 4.56 × 10⁻¹). These results align with previous findings in mouse and rat models, where ketamine similarly reduced the expression of IL-1β, IL-6, CCL2, and IL1-R2, reinforcing its role in attenuating inflammatory mediator production [48–50]. Additionally, research on spinal cord injury (SCI) in rats demonstrated that interventions reducing IL-6 and IL-1β levels, such as the administration of isosteviol sodium (STVNa), can lead to functional recovery, underscoring the importance of these pathways in central nervous system inflammation [51].

Notably, IFI27 (*q =* 8.65 x 10^-41^) showed the second largest shift in fold change between the post ketamine therapy timepoints. IFI27 displayed dynamic regulation, with initial suppression at FUP1 (*fc* =-24.5) transitioning to up-regulation at FUP2 (*fc* = 3.09). This controlled inflammatory response aligns with animal model and cell culture findings of ketamine’s broader role in mitigating pro-inflammatory states addressing PTSD-related immune dysregulation [52, 53]. IL-1β, another key pro-inflammatory cytokine, also exhibited persistent up-regulation at both FUP1 (*fc* = 1.3) and FUP2 (*fc* = 2.3), reflecting a shift toward controlled immune signalling. This up-regulation reflects ketamine’s ability to recalibrate immune signalling by maintaining a controlled pro-inflammatory response, rather than complete suppression, which is crucial for resolving chronic inflammation without compromising immune function [54]. In PTSD, these cytokines are frequently elevated, contributing to chronic neuroinflammation and the impairment of brain regions such as the prefrontal cortex and hippocampus [55, 56]. By targeting these central inflammatory nodes, ketamine has the potential to dampen hyperactive immune responses and inhibit the systemic and neural inflammation associated with PTSD.

### 4.2 Activation of Anti-Inflammatory Pathways and Therapeutic Targets

The initial increase of antimicrobial peptide signalling (z *=* 1.63) through CAMP (*q =* 1.9 x ^10-4^) and wound healing pathways (z *=* 1), COL5A2 (*q =* 2 x 10^-7^), CXCL8 (*q =* 6.69 x ^10-4^) and COL17A1 (*q =* 2.52 x 10^-3^) suggests that ketamine may support anti-inflammatory functions. These antibacterial responses highlight the protective role of ketamine in reducing the risk of secondary infections, which is particularly important for immunocompromised individuals.

The sustained activation of these pathways at FUP2 indicates that ketamine may not only reduces excessive inflammation but could also promote immune homeostasis and repair processes. This is evidenced by the activation of inflammatory suppression pathways, including SOCS1 (q = 1.38 × 10⁻) and SOCS3 (*q* = 0.0002). This also supports previous research on the suppression of SOCS/ STAT pathways in PTSD patients [57]. The activation of IL-10 signalling pathway (z *=* 2.82) in FUP2 also aligns with rat models where ketamine enhances anti-inflammatory signalling by increasing IL-10 activity, which helps counterbalance pro-inflammatory cytokines in PTSD [48, 58].

These pathways offer potential targets for further therapeutic development. For instance, consistently activated signalling pathways like IL-10 and genes like CAMP, CXCL8, IL-6 and IFI27 could serve as biomarkers for predicting treatment response and monitoring relapse risk by indicating inflammatory recurrence.

This study identified a significant increase in neutrophil degranulation (z = 6.0) at FUP2, and while neutrophils are primarily associated with acute immune responses, they also play a pivotal role in resolving inflammation and facilitating tissue repair [18]. Primed neutrophils actively secrete cytokines and inflammatory mediators, and can present antigens via MHC-II to stimulate T-cells [59]. Within this network, 41 molecules, including CEACAM6 (*q* = 2.36 × 10⁻ ²), CAMP (*q* = 6.61 × 10⁻ ²²), DEFA1 (*q* = 4.06 × 10⁻²¹), and RNASE3 (*q* = 8.44 × 10⁻ ²¹) exhibited sustained activation, suggesting that ketamine may prime the immune system to resolve inflammation over time. In previous *in vitro* studies ketamine has demonstrated to acutely suppress neutrophil degranulation, as an anti-inflammatory response, these results indicate an initial anti-inflammatory response but that inflammation may be returning to a pre ketamine state at FUP2 [60].

### 4.3 Sustained Immune Modulation

One of the study’s most significant findings is the sustained activation of immune-related pathways at FUP2, including interferon alpha/beta signalling (*z =* 4), IL-17 signalling pathway (*z =* 3.36), and cytokine storm signalling (*z =* 4.26). These results underscore ketamine’s long-term effects on immune regulation, extending beyond its short-term impact at FUP1. This aligns with previous evidence that ketamine modulates both central and peripheral immune responses, creating an environment conducive to long-term neuroinflammatory resolution [61].

Interferon alpha/beta signalling was significantly up-regulated at FUP2, a key pathway in antiviral defence and immune communication. The apparent activation of this pathway in this study via the up-regulation of interferons including IFI27, IFIT1 (*q =* 2.44 x 10^-37^), and IFIT3 (*q =* 1.14 x 10^-15^), alongside the activation of RSAD2 (*q =* 3.72 x 10^-28^) and USP18 (*q =* 1.68 x 10^-10^), suggests that ketamine may activate this signalling pathway a key component in the innate immune system [62]. These findings are consistent with pre-clinical findings of ketamine’s ability to recalibrate maladaptive immune responses and restore immune homeostasis [48, 63].

The IL-17 signalling pathway also plays a crucial role in immune modulation. This study identified 16 genes involved in IL-17 signalling, including CCL2 (*q =* 5.56 x 10^-16^), LCN2 (*q =* 1.5 x 10^-13^) and LIF (*q =* 1.65 x 10^-5^), which exhibited amplified activity at FUP2, reflecting a shift from innate immune responses to sustained adaptive immune regulation. These sustained effects are particularly relevant to addressing the chronic immune dysfunction observed in PTSD patients. The antimicrobial peptide pathway, which nearly doubled in activity from FUP1 to FUP2, and the long-term activation of cytokine storm signalling supports preclinical studies in which ketamine’s role in enhancing systemic defences, reducing chronic inflammation, and promoting resilience against infections [48, 64]. These pathways reflect the impact ketamine may have on recalibrating immune responses, transitioning from acute activation to long-term stabilisation.

### 4.4 Linking Neuroplasticity and Inflammation

Mouse models have indicated chronic inflammation in PTSD can impair synaptic plasticity, contributing to cognitive deficits and emotional dysregulation [65, 66]. By promoting pathways involved in tissue repair and immune resolution, ketamine may help restore the neural environment required for synaptic regeneration and neuroplasticity [67]. For instance, the threefold increase in wound healing pathway activity from FUP1 to FUP2 aligns with previous research on ketamine’s ability to promote neuronal repair, synaptic connectivity and antimicrobial peptide activity [63, 68]. Together these functions reflect a potential role for ketamine in reducing inflammation and enhancing neuroplasticity.

The macrophage activation pathway (*z =* 2.84) is also a significant component of immune modulation, with this study identifying 16 differentially expressed molecules following ketamine treatment for PTSD, included IL-1β, BPI (*q =* 1.14 x 10^-13^), CXCL10 (*q =* 1.85 x 10^-9^) and SOCS1, emphasising its dual role in inflammation resolution and neuroplasticity enhancement. The short-term response (FUP1) of this pathway showed moderate activation, marked by the up-regulation of pro-inflammatory genes such as IL-1β and CXCL8, reflecting an acute immune response necessary for initiating repair processes. By FUP2, the macrophage pathway demonstrated a transition toward sustained anti-inflammatory activity, characterised by the up-regulation of the IL-10 signalling pathway and HBEGF (*q =* 1.45 x 10^15^), which are associated with immune resolution, tissue repair, and synaptic remodelling. This temporal shift suggests that ketamine may prime macrophages for an initial immune response, followed by a reparative and regulatory phase, fostering wound healing and enhancing neuroplasticity. These findings have been replicated in earlier rat models and highlight the role of macrophage pathways in recalibrating inflammation [65, 69], which may address the chronic immune dysregulation commonly observed in PTSD [70]. Ketamine has also been found to promote astrocyte activity, which supports the repair and maintenance of the blood-brain barrier, helping to mitigate the harmful effects of neuroinflammation [71]. This reduction in cytokine activity and immune cell overstimulation could translate to improved neuroplasticity and better regulation of the HPA axis, both of which are disrupted in PTSD.

Additionally, the short-term response presents a stronger emphasis on antimicrobial activity and acute immune defence, while the sustained response integrates processes related to long-term inflammation and immune regulation, such as sustained lymphocyte activity and cellular infiltration by mononuclear leukocytes. These differences underscore the progression of ketamine’s effects, transitioning from rapid symptom alleviation to promoting long-term recovery and inflammation suppression and immune balance.

### 4.5 Addressing Chronic Inflammation and Relapse Prevention in PTSD

The sustained modulation of inflammatory pathways we found highlight ketamine’s ability to address the chronic low-grade inflammation characteristic of PTSD. At 1-week post-treatment, ketamine’s acute anti-inflammatory effects, with strong inhibition of key pro-inflammatory mediators such as CCL2, and IFI27, and bacterial proliferation in CAMP, highlight an immediate suppression of immune pathways. Furthermore, at 4-weeks post-treatment, a change to a more balanced response, with sustained activation of certain immune pathways including IL-17 signalling and interferon alpha beta signalling, alongside selective inhibition through IL-10 signalling, neutrophil degranulation and macrophage activation, suggests a shift from acute inflammation suppression. For instance, the neutrophil degranulation pathway showed a marked increase in activity from FUP1 to FUP2 (*z*: 3 to 6.008), suggesting that while there is an initial anti-inflammatory effect, inflammation may return to pre-ketamine levels over time. Similarly, the wound healing signalling pathway tripled in activity (*z*: 1 to 3.3), supporting *in vivo* previous findings where ketamine’s influence on reparative and neuroplastic processes is key for recovery from trauma-related neuronal damage [72]. These findings emphasize the progression of ketamine’s effects, shifting from rapid symptom relief and inflammation suppression to long-term immune regulation. While these sustained effects provide evidence for ketamine’s potential to prevent relapse by maintaining immune and inflammatory balance, the gradual return of some genes to expression levels comparative to baseline may suggest the need for maintenance dosing schedules or adjunct therapies to sustain these benefits. Together, they highlight ketamine’s long-term impact on inflammation and tissue repair as a crucial aspect of its therapeutic action, providing a molecular basis for its ability to prevent PTSD symptom recurrence and support sustained recovery.

### 4.6 Clinical Implications

As precision psychiatry research progresses, prominent developments in the field of neuroinflammation demonstrate its link to diseases, such as depression, PTSD, schizophrenia, bipolar disorder, and Alzheimer’s disease [73, 74]. Emerging evidence suggests that chronic low grade neuroinflammation plays a pivotal role in the pathophysiology of these conditions, mediated by overactive immune responses, cytokine imbalances, and microglial dysfunction [7, 75]. Existing approaches have identified biomarkers such as inflammatory cytokines IL-6 and TNF-α and neuroimaging markers in affected individuals [75, 76]. This study has identified sustained molecular changes targeting a reduction in pro-inflammatory pathways, immune remodulation, and promoting anti-inflammatory pathways observed up to 4 weeks post treatment that underscore ketamine’s potential as a long-term, therapeutic option for neuroinflammatory diseases.

Although these results are promising, several clinical questions remain unresolved, and one key challenge is relapse prevention. This study identified several genes that while differentially expressed in the short-term, began to revert toward baseline levels by FUP2. This suggests that a key application for ketamine’s therapeutic effects may include maintenance dosing, although the variability in individual responses, such as the dynamic regulation of IFI27, highlights the importance of risk-averse approaches in developing personalised schedules including dynamic feedback and safety protocols. Further, the long-term chronic misuse of ketamine has also been found to illicit memory deficits and inhibit cognitive function, especially in frequent non-prescribed ketamine users, through unregulated self-administration, and should be carefully considered when treating patients at risk of ketamine use disorder [77]. A recent rodent study administering a daily high dose (60mg/kg) of intraperitoneal ketamine was found to increase systemic inflammation potentially leading to cognitive disfunction [52]. Additional studies have also found that, dependant on the chronicity and dosage, ketamine may also inhibit neutrophil degranulation, thereby promoting inflammation in the long term [18].

Further research is needed into longer term treatment protocols to optimise ketamine’s efficacy and minimise variability in therapeutic outcomes. Repeated dosing regimens or maintenance treatments (e.g., weekly, bi-weekly or monthly infusions), or the use of poly-pharmacology by way of combining anti-inflammatory medications may be a promising avenue for further research as a way to safely sustain ketamine’s therapeutic effects over longer periods.

### 4.7 Limitations and Future Research

While the findings provide valuable insights, the study’s limitations must be acknowledged. The small sample size (*n =* 23) limits the generalisability of the results, and the four-week follow-up period may not capture the full trajectory of long-term effects or potential relapses. This analysis was also based on samples from an open-label (non-randomised, unblinded) clinical trial evaluating oral ketamine for PTSD treatment, which may have introduced bias in self-reported clinical assessments. Additionally, since participants continued their existing medications throughout the trial, potential interactions and effects between ketamine and other treatments, most notably the pharmacologic effect of anti-depressants cannot be excluded. Despite these limitations, the study identified numerous and statistically robust transcriptomic changes and their associated pathways, even after adjusting for multiple comparisons.

## 5 Conclusion

This study provides evidence of ketamine’s ability to modulate gene expression and canonical pathways in PTSD, with sustained effects observed up to four weeks post-treatment. These findings align with existing animal model research that identifies ketamine’s capacity to recalibrate immune responses and reduce neuroinflammation, both hallmarks of PTSD pathology. By modulating pathways such as cytokine storm signalling, neutrophil degranulation, IL-10 signalling and antimicrobial pathways, ketamine reduced inflammation while enhancing immune homeostasis. Additionally, ketamine’s effect on immune, inflammatory, and neuroplastic pathways offers a multifaceted approach to trauma recovery, having to potential to address the chronic inflammatory states that often underlie PTSD while simultaneously promoting long-term immune and neuroplastic stability.

In conclusion, this study provides the first longitudinal transcriptomic evidence that ketamine modulates immune function in PTSD, offering a novel framework for understanding its therapeutic effects beyond traditional neurotransmitter-based models. By integrating ketamine into precision psychiatry approaches, leveraging biomarkers for treatment response, and exploring combination therapies, we may be able to develop more effective, sustained, and personalised interventions for PTSD and other neuroinflammatory disorders. Future research should aim to include larger and more diverse cohorts, extended follow-up periods, and advanced multi-omic analyses incorporating neuroimaging to unravel the causal mechanisms underlying ketamine’s therapeutic effects.

## Author information

- National PTSD Research Centre, Thompson Institute, University of the Sunshine Coast (UniSC), Birtinya, QLD Australia Nathan J. Wellington, Ana P. Bouças, Bonnie L. Quigley, Adem T. Can
- School of Health, UniSC, Sippy Downs, QLD Australia. Anna V. Kuballa, Nathan J. Wellington
- Centre for Bioinnovation, UniSC, Sippy Downs, QLD Australia Anna V. Kuballa, Bonnie L. Quigley, Nathan J. Wellington
- Sunshine Coast Health Institute, Sunshine Coast Hospital and Health Service, Birtinya, QLD Australia Bonnie L. Quigley, Nathan J. Wellington
- Thompson Brain and Mind Healthcare, Maroochydore, QLD Australia Jim Lagopoulos, Megan Dutton

## Author Contributions

Article conceptualisation, N.W., and A.K.; methodology, N.W. and A.K.; formal analysis, N.W. and A.K.; writing—original draft and editing, N.W.; writing—review editing and supervision, A.K., review and editing, B.Q, A.B, A.C, M.D and J.L. All authors have read and agreed to the published version of the manuscript.

## Ethics Statement

Participants were recruited from the Oral Ketamine Trial on Post-Traumatic Stress Disorder (OKTOP; Prince Charles Hospital HREC Approval: HREC/18/QPCH/288, UniSC Ethics Approval: A181190, ANZ Clinical Trials Registry ACTRN12618001965291). Genetic analysis and reporting are registered under the Genetic Biomarkers of Ketamine on PTSD project (GBOK; Prince Charles Hospital HREC Approval: HREC/18/QPCH/288, UniSC Ethics Approval: S211655). Informed consent was obtained from patients over the age of 18 with written approval before commencement of the research.

## Data Availability

Illumina RNA-Seq 20 million sequence paired end reads, raw and processed data alongside deidentified metadata has been registered with the Gene Expression Omnibus (**GSE288174**) and support the findings of this study. Deidentified behavioural scale results are available on request although original copies are protected for participant privacy.

## Declarations

This manuscript will contribute to a Doctorate by Research (PhD) from the University of the Sunshine Coast for author N.W.

## Data Availability

Illumina RNA-Seq 20 million sequence paired end reads, raw and processed data alongside deidentified metadata has been registered with the Gene Expression Omnibus (GSE288174) and support the findings of this study. Deidentified behavioural scale results are available on request although original copies are protected for participant privacy.

## Acknowledgements

We would like to thank the Thompson Institute and the University of the Sunshine Coast for their support of molecular research into PTSD.

